# Dysregulated immune responses in COVID-19 patients correlating with disease severity and invasive oxygen requirements

**DOI:** 10.1101/2021.09.14.21263541

**Authors:** Paulina García-González, Fabián Tempio, Camila Fuentes, Consuelo Merino, Leonardo Vargas, Valeska Simon, Mirliana Ramirez-Pereira, Verónica Rojas, Eduardo Tobar, Glauben Landskron, Juan Pablo Araya, Mariela Navarrete, Carla Bastias, Rocío Tordecilla, Macarena A. Varas, Pablo Maturana, Andrés E. Marcoleta, Miguel L. Allende, Rodrigo Naves, Marcela A. Hermoso, Flavio Salazar-Onfray, Mercedes Lopez, María Rosa Bono, Fabiola Osorio

**Affiliations:** Laboratory of Immunology and Cellular Stress, Program of Immunology, Institute of Biomedical Sciences, Faculty of Medicine, Universidad de Chile, Santiago, Chile; Laboratory of Cancer Immunoregulation, Program of Immunology, Institute of Biomedical Sciences, Faculty of Medicine, Universidad de Chile, Santiago, Chile; Laboratory of Immunology, Biology Department, Faculty of Sciences, Universidad de Chile, Santiago, Chile; Nursing Department, Faculty of Medicine. Universidad de Chile, Santiago, Chile; Critical Care Unit, Department of Medicine, Hospital Clínico Universidad de Chile, Santiago, Chile; Laboratory of Innate Immunity, Program of Immunology, Institute of Biomedical Sciences, Faculty of Medicine, Universidad de Chile, Santiago, Chile; Laboratory of Antitumoral Immunology, Program of Immunology, Institute of Biomedical Sciences, Faculty of Medicine, Universidad de Chile, Santiago, Chile; Millennium Institute on Immunology and Immunotherapy, Faculty of Medicine, Universidad de Chile, Santiago, Chile; HIV Immunology and Allergies Unit, Department of Medicine. Hospital Clínico Universidad de Chile, Santiago, Chile; Integrative Microbiology Group, Biology Department, Faculty of Sciences, Universidad de Chile, Santiago, Chile; Center for Genome Regulation (CGR), Biology Department, Faculty of Sciences, Universidad de Chile, Santiago, Chile; Laboratory of Biochemistry and Molecular Biology, Biology Department, Faculty of Sciences, Universidad de Chile, Santiago, Chile; Laboratory of Neuroimmunology, Program of Immunology, Institute of Biomedical Sciences, Faculty of Medicine, Universidad de Chile, Santiago, Chile

**Keywords:** Severe COVID-19, Oxygen therapy, invasive mechanical ventilation, immunity

## Abstract

The prognosis of severe COVID-19 patients has motivated research communities to uncover mechanisms of SARS-CoV-2 pathogenesis also on a regional level. In this work, we aimed to understand the immunological dynamics of severe COVID-19 patients with different degrees of illness, and upon long-term recovery.

We analyzed immune cellular subsets and SARS-CoV-2-specific antibody isotypes of 66 COVID-19 patients admitted to the Hospital Clínico Universidad de Chile, which were categorized according to the WHO ten-point clinical progression score. These included 29 moderate patients (score 4-5) and 37 severe patients under either high flow oxygen nasal cannula (18 patients, score 6), or invasive mechanical ventilation (19 patients, score 7-9), plus 28 convalescent patients and 28 healthy controls. Furthermore, six severe patients that recovered from the disease were longitudinally followed over 300 days.

Our data indicate that severe COVID-19 patients display increased frequencies of plasmablasts, activated T cells and SARS-CoV-2-specific antibodies compared to moderate and convalescent patients. Remarkably, within the severe COVID-19 group, patients rapidly progressing into invasive mechanical ventilation show higher frequencies of plasmablasts, monocytes, eosinophils, Th1 cells and SARS-CoV-2-specific IgG than patients under high flow oxygen nasal cannula. These findings demonstrate that severe COVID-19 patients progressing into invasive mechanical ventilation show a distinctive type of immunity. In addition, patients that recover from severe COVID-19 begin to regain normal proportions of immune cells 100 days after hospital discharge and maintain high levels of SARS-CoV-2-specific IgG throughout the study, which is an indicative sign of immunological memory. Thus, this work can provide a useful benchmark for improvement of disease outcomes.

## Introduction

The pandemic of Severe Acute Respiratory Syndrome Coronavirus 2 (SARS-CoV-2), causing coronavirus disease 2019 (COVID-19), has led to millions of infections and deaths worldwide, with numbers continuing to increase (WHO dashboard information: https://covid19.who.int). As with other coronaviruses, SARS-CoV-2 is transmitted primarily via respiratory droplets, and upon infection, the median incubation period takes approximately 5-7 days. Disease outcomes range from asymptomatic and mild to more severe and critical courses with pneumonia, Acute Respiratory Distress Syndrome (ARDS), multiorgan failure, and considerable risk of fatality (1,2). The risk of developing severe disease has been associated with advanced age, comorbidities such as metabolic syndrome, lung and heart conditions and other factors such as poverty and social determinants in health (3–7).

In this context, symptomatic COVID-19 patients develop clinical manifestations within a 14-days window and can exhibit fever and dry cough, fatigue, anosmia, dyspnea, muscle and joint pain, headache, diarrhea and other symptoms (3,8,9). One factor that appears to be related to disease severity and outcome is the requirement for exogenous oxygen supplementation. Critically ill COVID-19 patients rapidly develop ARDS and require oxygen therapies. These therapies can be grouped into those that deliver high flow oxygen, such as high flow nasal cannula (HFNC), and those that require intubation, such as invasive mechanical ventilation (IMV)(10–12). The association between oxygen dependency, disease severity and mortality, is a parameter that the World Health Organization (WHO) considers for clinical classification of patients (13).

During this ongoing and evolving pandemic, rapid collaboration from scientists across the globe has begun to unravel the pathogenesis of COVID-19, leading to the definition of patient immune dynamics, clinical courses, and development of different therapeutic strategies to prevent disease progression. These studies have identified components of innate and adaptive immunity dysregulated in symptomatic adult patients (14–18), which include exacerbated inflammation characterized by neutrophilia, lymphopenia, T cell exhaustion and myeloid cell activation as hallmarks of severe outcomes (14,15,19– 24). However, whereas most studies have focused on the early stages of the disease, generally within the time surrounding symptom onset and patient hospitalization, fewer reports have addressed the dynamics between disease evolution and immune recovery spanning more extended periods (over three months) of study. Furthermore, the characterization of immune differences in severe COVID-19 patients with different degrees of illness has not been exhaustively studied.

Moreover, few studies have emerged from the Latin-American region, one of the most severely impacted by the COVID-19 pandemic. In this context, regional studies are essential considering the emergence of local SARS-CoV2 variants which may pose an increased risk to human health, and which may confer heterogeneity in clinical outcomes (7,25–28). Genomic surveillance programs in Chile have determined that, during the last year, the country has been successively impacted by most of the variants of concern (VOCs) defined by the WHO, including the B.1.1.7 (Alpha) SARS-CoV-2 variant, with increasing incidences of the P.1 variant (Gamma, which originated in the Amazonas’s state, Brazil) since January 2021, and more recently, Delta. In addition, there has been a recent surge of the C.37 variant (Lambda), variant of interest (VOI) first documented in Peru (https://auspice.cov2.cl/ncov/chile-global-2021-07-02; https://www.who.int/en/activities/tracking-SARS-CoV-2-variants/). Thus, a better understanding of immunity in response to SARS-CoV-2 at the regional view may offer important insights into disease management while contributing to obtaining a broader overview of how this new virus behaves under different geographic, ethnic, social, and public health conditions.

This one-year study assessed the innate and adaptive immune landscape of a heterogeneous cohort of 66 COVID-19 adult patients ranging from severe to moderate, who were admitted to the Hospital Clínico Universidad de Chile in Santiago, Chile. These patients were further classified according to the WHO Clinical Progression Scale (13) and were compared with 28 COVID-19 recovered/convalescent adult patients and 28 healthy controls. We report that severe patients who rapidly require invasive mechanical ventilation display an immunological profile typified by increased frequencies of plasmablasts, monocytes, eosinophils and Th1 equivalents, which distinguished this group from more stable severe COVID-19 patients treated with high flow nasal cannula. Finally, a small group of severe COVID-19 patients that recovered from the disease were longitudinally followed for ten months after having a positive PCR result for SARS-CoV-2. Our data show that these patients started to regain average proportions of key immune cell components dysregulated at the peak of the pathology including granulocyte composition, plasmablasts, and T cell activation, after three months of hospital discharge. These patients also maintained elevated titers of anti-Spike IgG throughout the study, which indicates the proper establishment of immunological memory. As such, these parameters offer a clear method for segregation between severe and recovered patients.

## Material and methods

### Ethics statement

This study was approved by the Institutional Review Boards at Hospital Clínico Universidad de Chile and at the Faculty of Medicine, Universidad de Chile (Protocol ID. Number 1151/20 and Protocol ID. Number N° 074-2020). All patients and healthy controls were required to understand the study and sign an informed consent.

### Study Design and recruitment

This study performed from July 2020 to August 2021, was designed to address the immunological response of COVID-19 patients. Twenty-eight healthy controls (Mean: 45 years; Interquartile Range (IQR): 26-58,5) were included in this study. Twenty-eight convalescent patients (Mean: 39 years; IQR: 29-52) were included in this study if they met the criteria of an actual positive PCR result for SARS-CoV-2 (Mean: 118,5 days; IQR:103,8-131,8) before the blood collection. The nursing team collected blood samples of healthy controls and convalescent patients. Sixty-six patients with confirmed SARS-CoV-2 infection by PCR were admitted to the Hospital Clínico Universidad de Chile between September 2020 and July 2021, were included in this study. A physician monitored COVID-19 patients, and COVID-19 disease severity scores were assigned according to the ten-point scale for COVID-19 trial endpoints of the WHO(13), which were matched to the review of electronic medical records of the patients. In this manner, 29 COVID-19 patients were scored with moderate disease status (Mean: 62 years; IQR: 51-69; WHO COVID-19 clinical scores 4-5) by presenting SARS-CoV-2 infection and requiring hospitalization without supplemental oxygen or requiring non-invasive supplemental oxygen. Thirty-seven COVID-19 patients were scored with severe disease status (Mean: 64 years; IQR: 57-71; WHO COVID-19 clinical scores 6-9) by requiring admission into intensive care unit (ICU) and requiring non-invasive ventilation or requiring invasive mechanical ventilation. All severe COVID-19 patients received corticoid treatment. The clinical data was stored using REDCap (Research Electronic Data Capture) software(29).

### Sample processing and cell isolation

Whole blood for flow cytometry analysis was collected in EDTA-coated vacutainers, and all blood samples were processed the same day as collection. Before PBMC isolation, 500ul of blood was aliquoted and stabilized with 25ul of Transfix (Cytomark) and stored at 4°C in the dark until staining. Blood and isolated PBMC aliquots (approximately 3×10^6^ cells) were stabilized using TransFix® (Cytomark) and stored for 5-7 days at 4°C before cell staining and acquisition for immunophenotyping.

### Isolation of PBMCs

Peripheral blood was diluted 1:1 in RPMI-1640 (Gibco) and layered into a SepMate tube (Stemcell Technologies) pre-loaded with 15ml of Lymphoprep (Stemcell Technologies). SepMate tubes were centrifuged for 20 minutes at 1200g and 20°C without brake and acceleration. The PBMC layer was collected, washed with RPMI for 15 minutes at 600g and 20°C, and treated with ACK lysis buffer (Thermofisher) for 10 minutes before dilution in RPMI for cell count and sample preparation for storage and staining. Approximately 3×10^6^ of isolated PBMC was resuspended in 1mL of RPMI and 5% of Transfix and stored at 4°C in the dark for 5-7 days until staining.

### Isolation of patient serum

Serum samples were collected after whole blood centrifugation at 400 g for 10 minutes at RT without brake. The undiluted serum was then transferred to 15 ml polypropylene conical tubes, aliquoted, and stored at -80 °C for subsequent analysis.

### ELISA

The ELISA protocol was previously adapted from the group of Kramer (30). Briefly, 96-well ELISA plates (MaxiSorp, Thermo Fisher) were coated overnight at 4°C with 50 µl per well of a 2 µg/ml solution of resuspended SARS-CoV-2 Spike protein (Recombinant SARS-CoV-2 S protein S1, Biolegend) on PBS. Then, the coating solution was removed, and the wells were blocked for one hour at room temperature with 150 µl of 3% skim milk prepared in PBS-0.1% Tween-20 (TPBS). After this period, the blocking solution was removed, and the sera to be tested were added. The serum samples to be tested were heated to 56 ° C for 1 hour before use to reduce the risk of residual virus. 100 µl per well of serial dilutions of the sera prepared in 1% skim milk in 0.1% TPBS were added and incubated for 2 hours at room temperature. The plates were then washed three times with 250 µl per well of 0.1% TPBS. Next, 100 µl per well of HRP-conjugated anti-human IgG (HRP Donkey anti-human IgG Clone: Poly24109) or anti-human IgA (Goat Anti-Human IgA alpha chain (HRP) ABCAM ab97215) secondary antibody diluted 1:10,000 in 0.1% TPB was added and incubated for 1 hour at room temperature. The plates were then washed three times with 250 µl of 0.1% TPBS, after which 50 µl of TMB substrate solution (TMB Substrate Reagent, BD Biosciences) were added per well to reveal the reaction which was then stopped after 10 min by adding 50 µl per well of 1M orthophosphoric acid. Optical density at 450 nm was measured on a Molecular Devices Emax ELISA plate reader. The background was established at a DO equal to 0.10. AUC values and data were analyzed using Prism 9 (GraphPad). Differences between groups were calculated using Kruskal-Wallis test and Dunn’s multiple comparison post-test.

### Flow cytometry

1×10^6^ PBMC or 100μl of transfixed-blood were used per patient depending on the staining panel. Before staining, both samples were left to acquire room temperature. PBMC staining was done in 96-well V bottom plates while direct blood staining was performed in 5-mL round-bottom polypropylene tubes. PBMC was washed and resuspended in 50μl of FACS buffer (PBS 1X, 1% FBS, 2mM EDTA) with Fc block (Biolegend) and incubated for 10 min in the dark at room temperature. After washing, the pellet was resuspended in 50μl of antibody staining mix and incubated for 30 min in the dark at room temperature. Cells were washed with 200uL of FACS at 600g for 5 min and resuspended in FACS buffer for acquisition. For granzyme B staining, after surface staining, cells were fixed for 20 min in the dark at room temperature with IC fixation buffer (Biolegend) and then washed twice with Perm Buffer at 600g for 5 min before intracellular staining with 50μL of granzyme B antibody dilution in Perm Buffer. Cells were incubated for 30 min in the dark at room temperature, washed with Perm Buffer, and resuspended in FACS buffer for acquisition.

For direct blood staining, 50μL of the corresponding antibody staining mix was added to 100ul of blood samples and incubated for 30 min in the dark at room temperature. Samples were then treated with 500μL of 1X EasyLyse Erythrocyte-Lysing Reagent (Agilent) diluted in DI water, vortexed, and incubated for 20 min in the dark at room temperature before data acquisition. All flow cytometry antibodies were purchased from Biolegend (San Diego, CA, USA). The following antibodies were used on a dilution 1/100: CD4 FITC (OKT4); CD138 PE (DL-101); CD56 PerCP/Cy5.5 (HCD56); CD123 APC (6H6); CD14 Alexa Fluor 700 (63D3); CD8 APC/Cy7 (SK1); CD38 BV421 (HIT2); CD11c B510 (3.9); CD16 605 (3G8); CD19 BV650 (HIB19); PD-1 BV711 (EH12.2H7); CCR7 PE (G043H7); CCR4 PerCP/Cy5.5 (L291H4); CD27 APC (M-T271); CXCR3 Alexa fluor 700 (G025H7); CD127 APC/Cy7 (A019D5); Granzyme B BV421 (QA18A28); CCR6 BV510 (G034E3); CXCR5 BV605 (J252D4); PD-1 BV711 (EH12.2H7); CD11b FITC (ICRF44); PD-L1 PE (29E.2A3); CD19 PE/Dazzle 594 (HIB19); CD86 BV650 (IT2.2); CD64 BV711 (10.1); CD24 PE/Dazzle 594 (ML5); IgM APC/Cy7 (MHM-88); CD38 BV421 (HIT2); IgG BV510 (M1310G05). Additional antibodies used on a 1/200 dilution were: CD3 PE/Dazzle 594 (UCHT1); CD45 PE/Cy7 (HI30); HLA-DR BV785 (L243); CD45RA BV650 (HI100); IgD FITC (IA6-2). Samples were acquired in a BD LSRFortessa X-20 with the FACS Diva Software.

Unsupervised flow cytometry analysis of CD45^+^ cells in PBMC samples was done using Uniform Manifold Approximation Projection (UMAP) along with the FlowSOM automated clustering tool of FlowJo Software. 50.000 CD45+ cells from representative samples of each condition studied were concatenated and dimensionality reduction was assessed using the UMAP plugin from FlowJo (12 nearest neighbors, 0.5 minimun distance). For clustering visualization, we used the FlowSOM plugin (9 metaclusters). Parameters considered in the analyses include CD4 FITC, CD138 PE, CD3 PE/Dazzle 594, CD123 APC, CD14 Alexa Fluor 700, CD8 APC/Cy7, CD38 BV421, CD11c BV510, CD16 BV605, CD19 BV650, HLA-DR BV786.

### Statistical analysis

Statistical analyses were performed using Prism 8 software (GraphPad). Multiple group comparisons were calculated using non-parametric Kruskal Wallis test with Dunn’s multiple comparison post-test. Comparisons between two groups were assessed using Mann-Whitney t-tests. Results are shown as individual data with mean ± SEM for each group.

## Results

To better understand the different immune signatures of COVID-19 patients, we devised a 14-parameter flow cytometry strategy to stratify patients in terms of innate immune cell composition, lymphocyte populations, activated/exhausted CD4^+^ and CD8^+^ T cells, and B cell/plasmablasts, following a methodology adapted from reported work (14,16,19). This protocol was applied to whole blood (for analysis of granulocytes including neutrophils and eosinophils) or to peripheral blood mononuclear cells (PBMC, for analysis of lymphocytes and monocytes) (Scheme of the study is depicted in Supp. Fig.1 and gating strategy is depicted in Supp. Fig. 2). We also measured serum IgM, IgG, and IgA antibodies against SARS-CoV2 spike (S) protein and included a longitudinal follow-up study of 6 patients admitted to the Intensive Care Unit (ICU) and monitored up to 300 days after hospital discharge.

To classify COVID-19 patients according to disease severity, we employed the WHO ten-point clinical progression scale that provides standard outcome measures for COVID-19 studies (13). According to this scale, our study included 29 patients categorized as ‘Hospitalized Moderate disease’ (referred to as “M’’), which consisted of 5 patients with a score of 4 (hospitalized without oxygen therapy) and 24 patients with a score of 5 (oxygen by mask/nasal prongs). We also recruited 37 patients categorized as ‘Hospitalized Severe Disease ‘patients (referred to as “S”), consisting of 18 patients with a score of 6 (hospitalized with HFNC), 5 patients with a score of 7 (hospitalized with intubation and mechanical ventilation), 9 patients with score 8 (hospitalized with intubation, mechanical ventilation, PaO2:FiO2 < 150 or vasopressor treatment), and 5 patients with score 9 (hospitalized with intubation, mechanical ventilation, PaO2:FiO2 < 150 and vasopressor dialysis) (Table 1). This study also included a group of 28 convalescent COVID-19 patients who went through SARS-CoV2 infection 118,5 days (Median: 118,5 days; IQR: 103,8-131,8) before sample collection, according to the date of a positive SARS-CoV2 PCR. In our cohort, patients have a body mass index (BMI) of 28,6 (IQR: 22,5-33), 46,3%, have moderate hypertension and 15,8% have diabetes. We did not find significant differences in the BMI of moderate and severe patients (Table 1). However, among the severe patient group, we found a significantly higher number of individuals with moderate arterial hypertension compared to the moderate patient group. Three moderate and 5 severe COVID-19 patients died during the time of the study.

**Table 1.** Clinical Characteristics. Data are listed as median (IQR)

|  | Moderate (29) | Severe (37) | Convalescent (28) | All Patients (94) |
| --- | --- | --- | --- | --- |
| **Age** | 62 (51-69) | 63 (57-71) | 39 (29-52) | 60 (44-68) |
| **Sex** |  |  |  |  |
| Male | 11 (37,9%) | 22 (60%) | 7 (25%) | 40 (42,6%) |
| Female | 18 (62,1%) | 15 (40%) | 21 (75%) | 54 (57,4%) |
| **Body Mass Index** | 27,5 (23,7-28,8) | 29,6 (25,6-33) | 29,7 (22,4-31,7) | 28,6 (22,5-33) |
| **Time between PCR confirmation and first blood sample (days)** | 3 (1-10,3) | 3 (1-7) | 118,5 (103,8-131,8) | 3 (1-8,3) |
| **Hospitalization (days)** | 12 (6-16) | 23 (16-32) | - | 17 (11,3-25) |
| **Deceased** | 3/29 (10,3%) | 6/37 (16,2%) | - | 9/94 (9,5%) |
| **Comorbidity** |  |  |  |  |
| Diabetes | 3/29 (10,3%) | 7/37 (18,9%) | 5/28 (17,9%) | 15/94 (16%) |
| Arterial Hypertension | 10/29 (34,5%) | 24/37 (64,9%) | 10/28 (35,7%) | 44/94 (46,8%) |
| Coronary Heart disease | 2/29 (6,9%) | 3/37 (8,1%) | 3/28 (10,7%) | 8/94 (8,5%) |
| Obesity | 0/29 (0%) | 3/37 (8,1%) | 1/28 (3,6%) | 4/94 (4,3%) |
| Hypothyroidism | 1/29 (3,4%) | 0/38 (0%) | 2/28 (7,1%) | 3/94 (3,2%) |
| Malignancy | 1/29 (3,4%) | 1/38 (2,6%) | 1/28 (3,6%) | 3/94 (3,2%) |

| **WHO Score** | **Oxygen therapy** | **Nº Patients** |
| --- | --- | --- |
| Moderate 4 | Hospitalised, no oxygen therapy | 5 |
| Moderate 5 | Hospitalised, oxygen by mask or nasal prongs | 24 |
| Severe 6 | Hospitalised, oxygen by non-invasive ventilation or high flow | 18 |
| Severe 7 | Intubation and mechanical ventilation, SpO2/FiO2 ≥150 or SpO2/FiO2 ≥200 | 5 |
| Severe 8 | Mechanical ventilation SpO2/FiO2 <150 (SpO2/FiO2 <200) or vasopressors | 9 |
| Severe 9 | Mechanical ventilation SpO2/FiO2 <150 and vasopressors, dialysis, or ECMO | 5 |

Finally, our study includes 28 healthy controls (HC) matched by age and comorbidities. A positive SARS-CoV-2 PCR confirmed all COVID-19 patients before the analysis, and a negative result verified the absence of infection in healthy controls.

### Innate immune cell analysis as indicators of COVID-19 severity

We analyzed the immune subset composition of moderate, severe, and convalescent COVID-19 patients and healthy controls to identify significant discriminators of the pathology (gating strategy depicted in Supp Fig 2). In total blood, analysis of innate immune cells showed a marked presence of polymorphonuclear leukocytes (PMNs) and a clear increase in neutrophils in moderate and severe patients, which segregated them from healthy controls as reported (14,31), and also from convalescent patients (Fig 1a). Even though the frequencies of eosinophils oscillated within a reduced range, there was a significant reduction of these cells in moderate and severe patients compared to healthy controls and convalescent patients (Fig 1b). Furthermore, as reported(14), basophil frequency was drastically reduced in moderate and severe patients compared to healthy controls but also compared to convalescent individuals, suggesting that alterations in granulocyte populations occur during the onset of pathology (Fig 1c). We also observed significant reductions in plasmacytoid dendritic cell (pDCs) frequency in moderate and severe patients compared to healthy controls and convalescent patients (Fig 1d). Interestingly, even though monocyte frequency was not altered among COVID-19 patients (Supp Fig 3a, Fig 1e), the frequency of intermediate monocytes in moderate, severe, and convalescent patients was increased compared to healthy controls (Fig 1e), suggesting that the pathology re-shapes this myeloid cell repertoire even in patients that have recovered from the disease. Accordingly, non-conventional monocyte frequency was decreased in severe and convalescent patients (Fig 1e). Along these lines, severe COVID-19 patients downregulated expression of HLA-DR in all monocyte subtypes compared to healthy controls (Fig 1f, Supp Fig 3b), but convalescent patients showed expression of the molecule that was comparable to that of healthy controls. This data is accompanied by decreased expression of the costimulatory molecule CD86 in monocyte subtypes from severe patients compared to healthy controls (Fig 1g).

**Figure 1.**
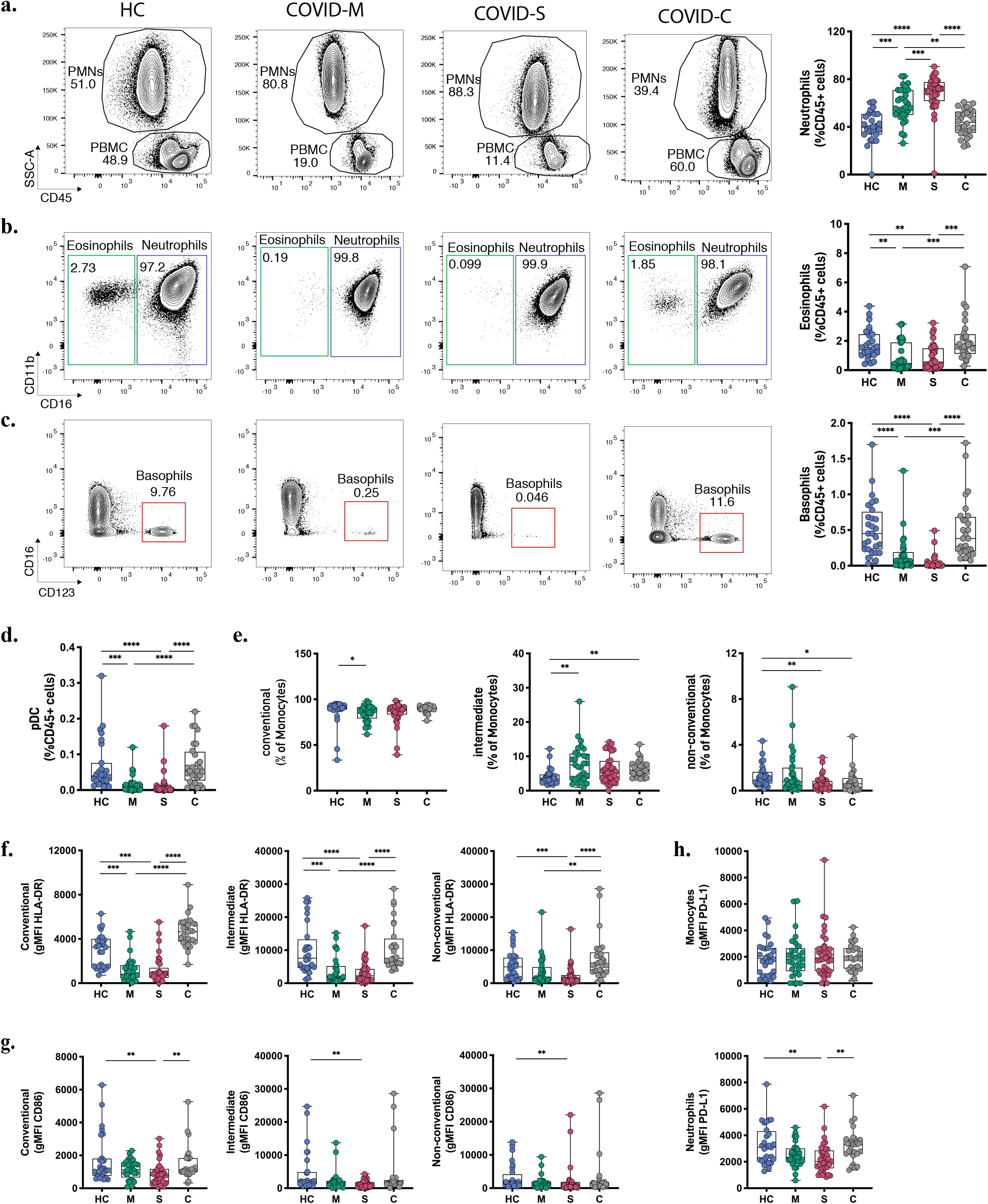
Innate Immune profile of COVID-19 patients. Frequency and phenotype of granulocytes and other innate immune populations were assessed through multiparametric flow cytometry analysis of peripheral blood samples of COVID-19 patients with moderate and severe disease, convalescent COVID-19 patients and healthy controls. **A-C**. Representative flow cytometry plots and frequency of granulocyte populations calculated within CD45^+^ leukocytes. **A**. Polymorphonuclear cells (PMN) and peripheral blood cells (PBMC) gates within CD45^+^ cells for each subject studied. **B-C**. Neutrophils, eosinophils and basophils frequency was assessed for each donor. FACS plots show percentages of each population within the PMN gate (neutrophils and eosinophils) or the PBMC gate (basophils) whilst box plots display cell frequency inside the CD45^+^ population. **D**. Plasmacytoid dendritic cell (pDCs) frequency within the CD45+ population. **E-G**. Frequency of conventional, intermediate and non-conventional monocytes subsets found in peripheral blood samples (**E**) and the mean fluorescence intensity of HLA-DR (**F**) and CD86 (**G**) within the different monocyte subsets. **H**. PD-L1 expression levels in neutrophile and monocyte populations of COVID-19 patients and healthy controls. HC = Healthy controls, n = 28; M = COVID-Moderate, n = 29; S = COVID-Severe, n = 37; C = COVID-Convalescent, n = 28. Differences between groups were calculated using Kruskal-Wallis test and Dunn’s multiple comparison post-test (*p = 0,05; **p = 0,01; ***p = 0,001, ***p = 0,0001).

Finally, we measured surface expression levels (through the Mean Fluorescence Intensity, MFI) of PD-L1, an immunoregulatory molecule with reported roles in myeloid cells in COVID-19 (32). Our data showed that neutrophils from severe patients have decreased PD-L1 expression compared to healthy controls and convalescent patients (Fig 1h, Supp Fig 3c). Altogether, these data indicate that severe COVID-19 patients show a dysregulated innate immune cell balance, evidenced at the level of granulocyte composition and in the repertoire and activation status of monocytes.

### Adaptive immune signatures of COVID-19 patients

Next, we sought to characterize the dynamics of lymphocytes in COVID-19 patients throughout different stages of the pathology. Data depicted in Figure 2a indicates that B cell percentages among CD45^+^ cells are markedly decreased in moderate and severe patients but are restored in convalescent individuals, in agreement with the notion that COVID-19 patients with ongoing infection show lymphopenia (22,24). Notably, transitional B cells, representing an intermediate developmental stage between immature B cells in the bone marrow and mature peripheral B cells (14), were significantly increased in moderate, severe and convalescent patients compared to healthy controls, which suggests that convalescent patients show long term cellular changes associated to the infection (Supp Fig 4a). Furthermore, although we did not observe significant changes in the frequency of näive or IgD-CD27-B cells, we found a decrease in B cell memory switch in severe and moderate patients compared to healthy controls and convalescent patients (Supp Fig 4b). Remarkably, plasmablasts, which are dividing antibody-secreting cells circulating in the blood (33) were detected in COVID-19 patients in a pattern that correlated with disease severity (Fig 2b-c). As such, these data prompted us to investigate the antibody responses in COVID-19 patients (Fig 2d). To this end, we performed ELISAs to detect antibodies specific against the Spike (S) protein of SARS-CoV-2. These assays indicated that moderate and severe COVID-19 patients produced detectable levels of S-specific IgM, IgG and IgA antibodies, suggesting that SARS-CoV-2 infection triggers universal production of antibody isotypes against the virus. In addition, convalescent patients expressed high levels of S-specific IgG but not IgM antibodies, agreeing with a temporal resolution of the infection and with the generation of immune memory (Fig 2d). Altogether, these data indicate that plasmablasts and the presence of antibody isotypes are hallmarks of COVID-19 patients, segregating these groups from patients that have resolved the infection.

**Figure 2.**
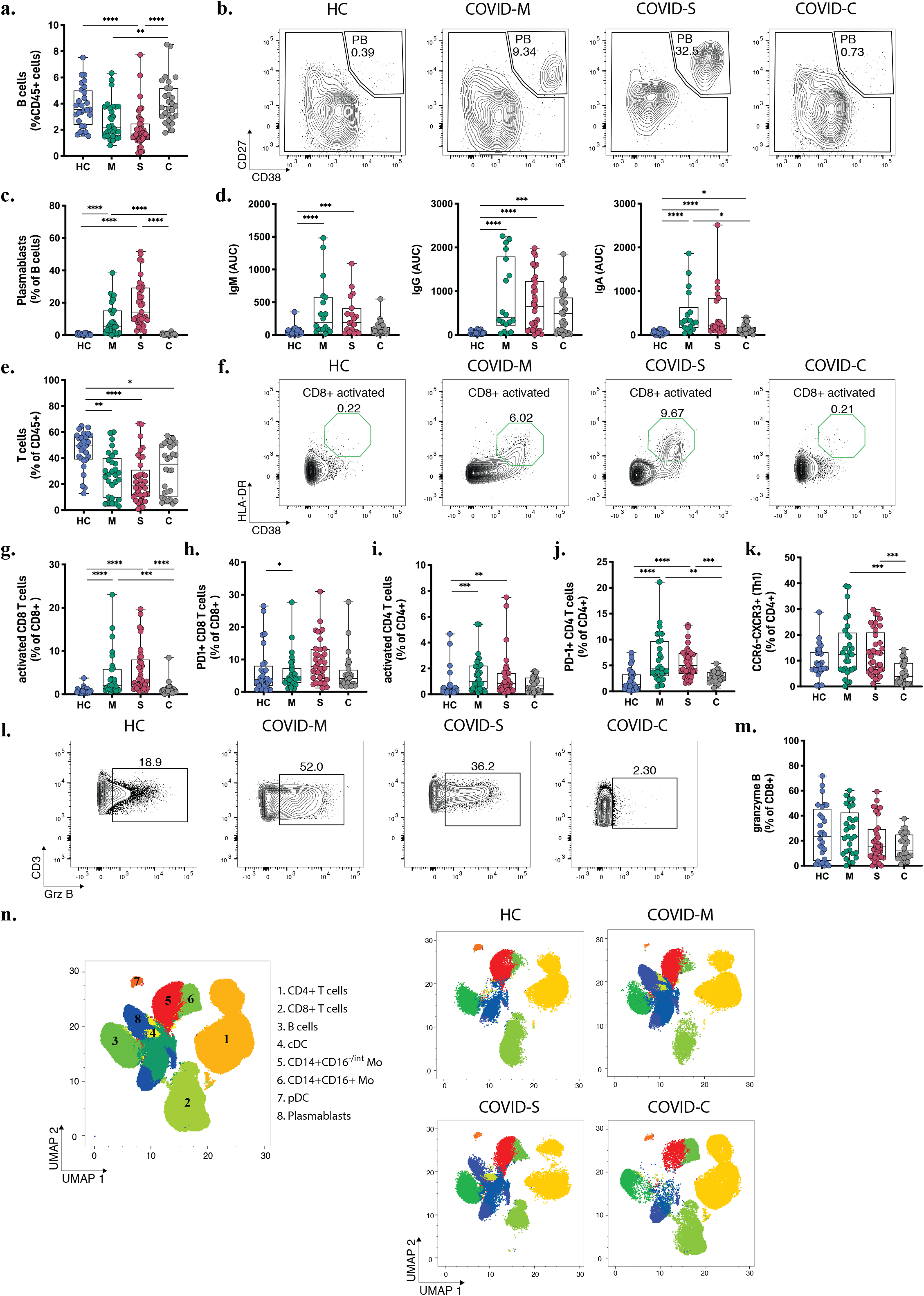
Immune profiling of adaptive immune responses in COVID-19 patients. B and T cell populations were analyzed through multiparametric flow cytometry in peripheral blood samples of COVID-19 patients with moderate and severe disease, convalescent COVID-19 patients and healthy controls. **A**. Frequency of B cells within CD45^+^ leukocytes. **B-C**. Representative plots and percentages of plasmablasts populations. **D**. SARS-CoV-2 Spike protein-specific immunoglobulin levels measured in plasma from COVID-19 patients and healthy controls. **E**. Frequency of T cells within CD45^+^ leukocytes in peripheral blood. **F-G**. Representative plots and frequency of activated (F-G) and exhausted (H) populations within the CD8 T cell compartment. **I-K**. Frequency of activated (I) exhausted (J) and Th1 equivalents (CCR6^-^CXCR5^+^) populations within the CD4 T cell compartment. **L-M**. Representative plots and frequency rates of granzyme B+ CD8 T cells. **N**. UMAP of flow cytometry data of main leukocyte populations present in PBMC samples with representative images for moderate, severe, and convalescent COVID-19 patients in addition to healthy controls. Key numbers for cell subsets are indicated in the concatenated image. HC = Healthy controls, n = 28; M = COVID-Moderate, n = 29; S = COVID-Severe, n = 37; C = COVID-Convalescent, n = 28. Differences between groups were calculated using Kruskal-Wallis test and Dunn’s multiple comparison post-test (*p = 0,05; **p = 0,01; ***p = 0,001, ****p = 0,0001).

Next, we analyzed T cell dynamics in SARS-CoV-2 infected individuals. In agreement with reported work (14,22,24), moderate and severe COVID-19 patients showed decreased T cell frequencies in a severity-related manner (Fig 2e). Interestingly, although convalescent patients restore B cell frequencies to levels comparable to those observed in healthy controls (Fig 2a), these individuals do not restore T cell populations to normal levels (Fig 2e), suggesting that lymphopenia recovery occurs at different timings for B cells and T cells. We next determined the activated/exhausted status of CD4^+^ and CD8^+^ T cells (Fig 2f-h) and observed that patients with moderate and severe COVID-19 show increased accumulation of activated CD8^+^ T cells, which distinguished these cohorts from healthy controls and convalescent individuals (Fig 2f-h). In addition, CD8^+^ T cells expressing the exhaustion marker PD-1 accumulated in moderate patients and there was a clear, albeit not significant, trend towards higher frequencies in severe COVID-19 patients (Fig 2h). On the other hand, analysis of activated CD4^+^ T cells showed a similar tendency, with high frequencies in moderate and severe patients compared to healthy controls (Fig 2i), whereas CD4^+^PD-1^+^ T cells segregated moderate and severe patients from healthy controls and convalescent individuals (Fig 2j). Furthermore, using chemokine receptors as correlates of T helper subsets (14), we found that the frequency of Th1 equivalents (CCR6^+^CXCR3^+^) was unaltered between healthy controls, moderate and severe COVID-19 patients, but it was increased compared to convalescent individuals (Fig 2k). Frequencies of Th17 equivalents (CCR6^+^CXCR3^-^) remained unaltered among groups (Supp Fig 2c). Finally, we investigated if the production of granzyme B, a primary cytotoxic mediator, distinguished COVID-19 patients depending on the severity of the disease (Fig 2l-m). Our data indicate that expression of granzyme B in CD8^+^ T cells was not different between healthy controls, moderate and severe patients suggesting that this molecule does not discriminate between different degrees of SARS-CoV2 infection. However, we noticed that convalescent individuals express markedly low molecule levels, possibly reflecting resolution from an effector immune response (Fig 2l-m). In summary, these data suggest that hospitalized COVID-19 patients show a T cell signature typified by an accumulation of activated CD8^+^ and CD4^+^ T cells and CD4^+^ PD-1^+^ T cells. Finally, to obtain a broad overview of immune cell dynamics of COVID-19 patients, we devised an unsupervised flow cytometry analysis of CD45^+^ cells to cover representative innate and adaptive cells present in the PBMC. Data were visualized using a Uniform Manifold Approximation Projection (UMAP) (Fig 2n). PBMCs were grouped into populations by FlowSOM automated clustering tool of FlowJo Software for unbiased identification of leukocytes in healthy controls, moderate, severe, and convalescent patients (Fig 2n). This analysis identified 8 main clusters corresponding to CD4^+^ T cells (cluster 1, CD3^+^CD4^+^); CD8^+^ T cells (cluster 2, CD3^+^CD8^+^), B cells (cluster 3, CD3^-^CD19^+^), cDCs (cluster 4, CD3^-^HLA-DR^+^CD14^-^CD11c^+^), monocytes (Cluster 5, CD3^-^CD14^+^CD16^-/int^ and cluster 6, CD3^-^CD14^+^CD16^+^), pDCs (cluster 7, CD3^-^HLA-DR^+^CD14^-^CD123^+^) and plasmablasts (cluster 8, CD3^-^CD19^+^CD138^-^CD38^+^) (Fig 2n). We observed changes in the composition of pDCs, CD8^+^ T cells and CD4^+^ T cells, along with the emergence of plasmablasts in moderate and severe COVID-19 patients, a feature no longer observed in convalescent patients. Altogether, this data confirms our previous findings obtained through manual gating analysis (Fig 1d, 2c, 2e) and supports the conclusion that hospitalized COVID-19 patients display marked changes in the composition of pDCs, T cells and plasmablasts.

### Immune cell signatures associated with severe COVID-19 states

Our data indicate that severe COVID-19 patients show significant changes in subset composition of innate and adaptive immune cells. These findings prompted us to investigate whether there was a correlation between immune cell frequency and disease severity in this group of patients. To this end, we subclassified severe COVID-19 patients according to the World Health Organization (WHO) ten-point clinical progression scale for COVID-19 patients and analyzed immune parameters associated with each group. Eighteen severe patients categorized in score 6 and corresponding to hospitalized patients with HFNC were compared to 19 severe patients categorized in scores 7-9, which corresponds to hospitalized patients with intubation and mechanical ventilation with or without vasopressor use. Data in Figure 3 indicates that several immune cell types including B cells, CD4^+^ and CD8^+^ T cells, activated and PD-1^+^ CD4^+^ and CD8^+^ T cells, neutrophils, DCs, and basophils did not discriminate between score 6 and score 7-9 patients. In addition, D-dimer concentration in plasma was similarly elevated in both groups, concordant with the clinical severity of the disease (Fig 3a). However, specific immune cells including plasmablasts, monocytes, eosinophils, and Th1 equivalents, were increased in patients with higher clinical scores (Fig 3b-e). Remarkably, the concentration of S-specific IgG also differentiated patients under HFNC treatment versus invasive ventilated patients, with higher antibody titers in patients with higher severity rank (Fig 3b). Conversely, granzyme B expression was reduced in patients with mechanical ventilation compared to HFNC patients (Fig 3d). Altogether, these data indicate that patients that required invasive mechanical ventilation display an exacerbated innate and adaptive immune response typified by increased frequencies of plasmablasts, monocytes, Th1 equivalents, and S-specific IgG, which may provide relevant information regarding the progression of severe COVID-19 pathology.

**Figure 3.**
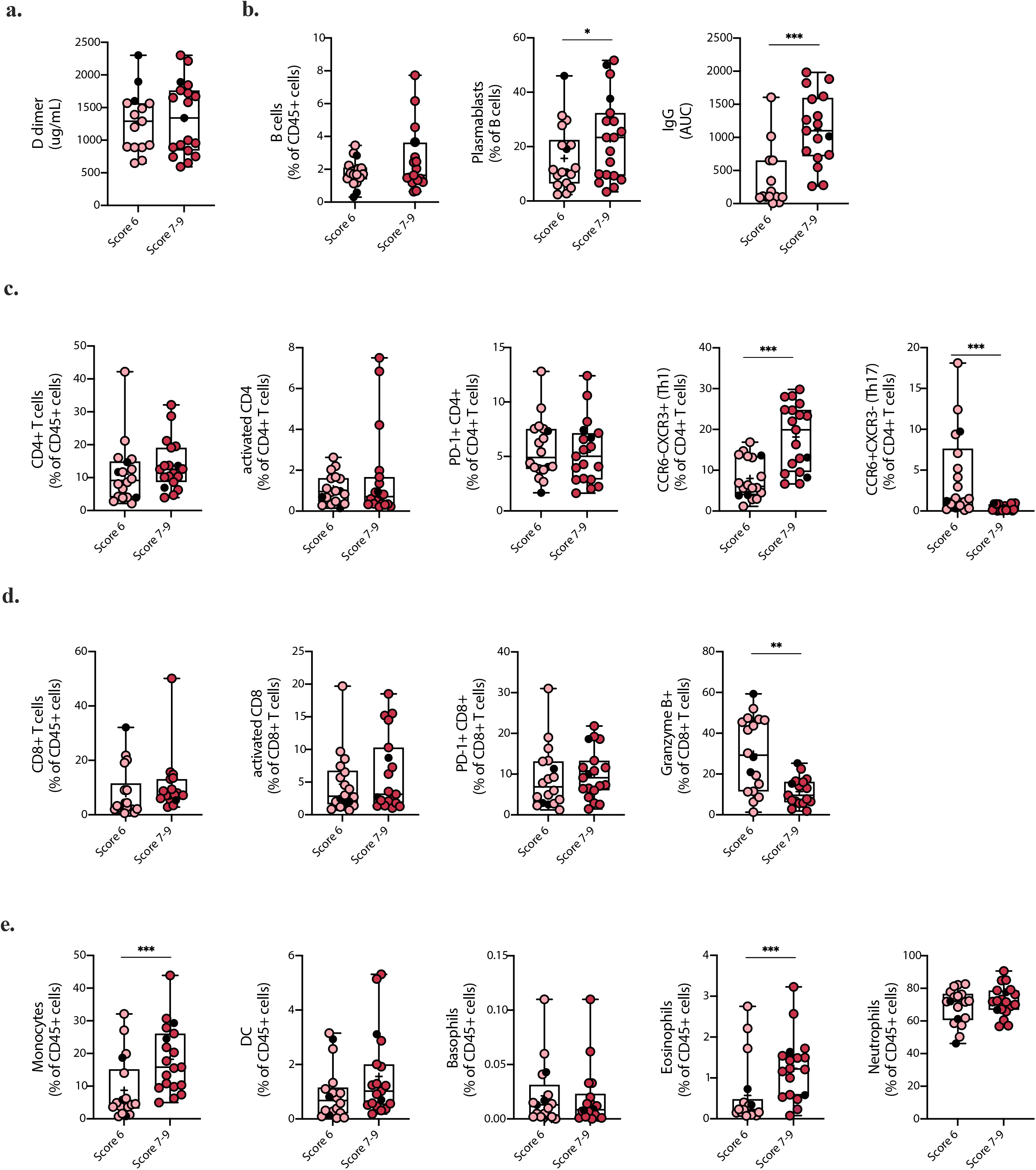
Clinical parameters and differences in the immune cell landscape between Severe COVID-19 patients with different severity scores. Changes among key clinical parameters and main immune cell populations in peripheral blood of COVID-19 patients with severe SARS-CoV-2 infection (COVID-S) were assessed within patients with different severity scores. **A**. D-dimer plasma concentrations. **B**. Humoral response of COVID-S patients with different severity scores. B cell frequency on CD45^+^ cells, plasmablasts percentage within the B cell population and Immunoglobulin G (IgG) concentration in plasma. **C-D**. Cellular immune response in COVID-S patients with different severity scores. **C**. CD4^+^ T cell frequency within the CD45^+^ compartment, activated and PD-1^+^ CD4^+^ T cell populations and Th1 equivalents percentage of CD4^+^ T cells. **D**. CD8^+^ T cell frequency within the CD45^+^ compartment, activated and PD-1^+^ CD8^+^ T cell populations and granzyme B^+^ CD8^+^ T cells. **E**. Myeloid cell and granulocyte compartment analysis from peripheral blood of COVID-19 severe patients classified by disease score. Graphs show frequency of monocytes, dendritic cells (DC), basophils, eosinophils and neutrophils within CD45^+^ cells. Score 6 = 18; Score 7-9 = 19. Black circles indicate deceased patients. Differences between groups were calculated using Mann Whitney tests (*p = 0,05; **p = 0,01; ***p = 0,001). Data mean is shown by a + on each box plot.

### Longitudinal analysis of severe COVID-19 patients on their transition to recovery

To better understand how the immune system of severe COVID-19 patients recovers after infection, we carried out a longitudinal analysis of 6 severe patients, which were followed starting from ICU hospitalization up to 280 days after clinical discharge (200-300 days after SARS-CoV-2 PCR confirmation). Samples were collected on 5 different time points regarding the date of the positive PCR for SARS-CoV2: T1 (Timepoint 1: Median: 1 day; IQR: 1-7), T2 (Timepoint 2: Median: 6,5 days; IQR: 6-11), T3 (Timepoint 3: Median: 12,5 days; IQR 10-16), R1 (Recovery point 1; Median: 119,5 days; IQR 112-130 after positive SARS-CoV-2 PCR and; Median: 103 days; IQR: 92-108 after hospital discharge) and R2 (Recovery point 2; Median: 300 days; IQR: 294-310 after positive SARS-CoV-2 PCR and; Median: 284 days; IQR: 269-294 after hospital discharge) (Scheme depicted in Fig 4a). Data depicted in Fig 4b-d shows that immune cells correlating with disease severity such as neutrophils and plasmablasts are markedly reduced in the R1 and R2 time points, suggesting a decline in effector immune cell types in recovered patients. Accordingly, basophils, reduced in severe patients with ongoing infection, were increasingly restored in the R1-R2 time points, along with a recovery of CD8^+^ and CD4^+^ T cells and B cell populations (Fig 4b-c). These data indicate a reversal of the pan-lymphocytopenia reported for severe COVID-19 patients (19). Remarkably, plasma levels of S-specific IgG remained elevated 300 days after infection (Fig 4c), confirming the notion that COVID-19 patients generate a durable immunological memory (34). In contrast, we also measured the trajectory of S-specific IgA and found that titers of the isotype decline over time (Fig 4c). Finally, we also observed a constant decrease in the frequencies of activated and PD-1^+^ CD8^+^ and CD4^+^ T cells during R1-R2 time points. Altogether, these data indicate that severe COVID-19 patients that can resolve SARS-CoV-2 infection show signs of restoration of specific immune cell compartments on a trajectory that resembles transition to homeostasis and generation of immunological memory.

**Figure 4.**
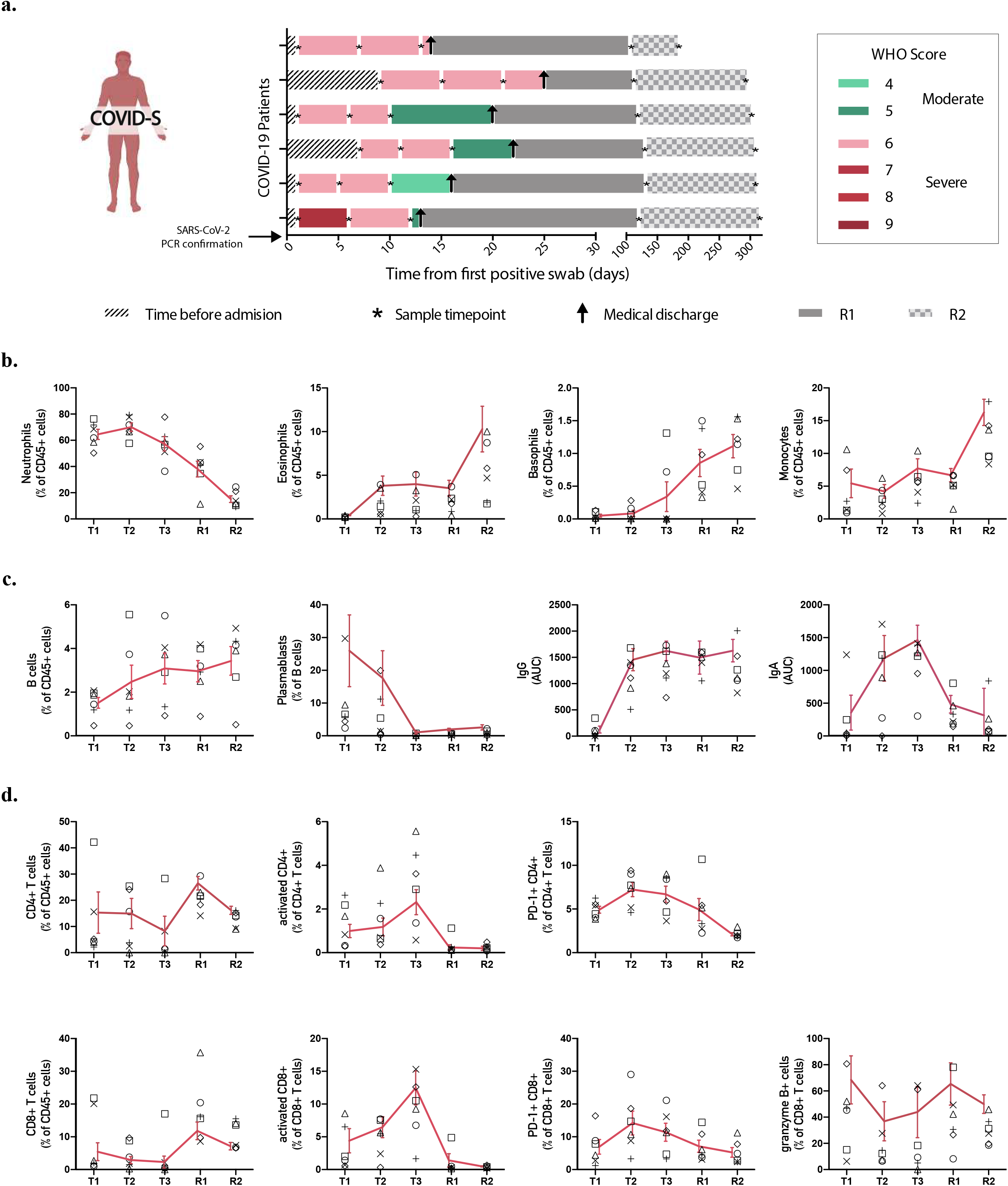
Longitudinal analysis of immune populations in patients with severe SARS-CoV-2 infection. Main immune cell populations affected in severe COVID-19 patients were analyzed over time at different intervals until patient recovery. **A**. Follow-up diagram of patients with severe COVID-19 disease. Peripheral blood samples from 6 patients with severe SARS-CoV-2 infection were collected at 3 different time points after ICU admission and SARS-CoV-2 PCR confirmation (T1, T2 and T3), plus two additional samples after recovery and clinical discharge (R1 and R2). The first sample (T1) was taken upon ICU admission and positive SARS-CoV-2 PCR, while second (T2) and third (T3) blood samples were taken 5-7 and 9-13 days after, correspondingly. R1 samples for follow-ups were taken once the patient had recovered and been released from the hospital, between 1 and 3 months after ICU admission. R2 samples were taken 6 months after patient recovery and clinical discharge. **B**. Changes in frequency of innate immune populations during disease evolution and recovery. **C-D**. Changes of adaptive immune components throughout time. Graphs show individual data for each patient and the mean ± SEM per group connected by a red solid line.

## Discussion

Although SARS-CoV2 infection generally leads to a mild disease in a large proportion of infected individuals, 5-15% of COVID-19 patients develop a severe pathology that progresses to pneumonia and respiratory failure (35–38). In this context, the immune cell landscape of severe COVID-19 patients is reported to be dysregulated (14,19,31,32), and detailed insights on the cellular dynamics of severe COVID-19 patients are urgently needed to identify potential disease intervention points. In this work, we analyzed immune cell dynamics of moderate, severe, and convalescent COVID-19 patients and further investigated the dynamics of severe patients in terms of oxygen dependence and long-term trajectories of ICU patients during the acute symptomatic stage and after recovery.

Previous studies have shown dysregulation of innate and adaptive immune cell compartments in patients with moderate, severe, and convalescent COVID-19 (14,19,22). Our data revealed statistically significant changes in immune cell subset composition between severe and convalescent patients, including neutrophils, basophils, eosinophils, pDCs, B cells, plasmablasts, activated CD8^+^ and CD4^+^ T cells and Th1 equivalents, providing a complete description of the dynamics of acute COVID-19 responses. In addition, we detected alterations in the composition and function of monocytes in severe COVID-19 patients with an abundance of HLA-DR low monocytes, which indicate dysfunctional monocyte function (17,32). Furthermore, even though single-cell transcriptome studies revealed an upregulation of PD-L1 expression in neutrophils from severe COVID-19 patients (32), our data on surface PD-L1 expression in neutrophils showed an opposite effect, indicating that additional studies should be conducted to clarify this issue. In this context, multiparametric flow cytometry analysis for COVID-19 patients’ immunophenotyping does not necessarily correlate with changes observed at the transcriptomic level.

As per adaptive immune parameters, the frequencies of plasmablasts and activated CD8^+^ and CD4^+^ T cells were increased in severe COVID-19 patients. Additionally, we also found higher frequency of PD-1^+^ CD8^+^ and CD4^+^ T cells, suggestive of potential T cell exhaustion. As expected, antibody data indicated that moderate and severe COVID-19 patients produce S-specific IgM, IgG, and IgA. However, in contrast to previous observations (19,20), our data show that convalescent patients no longer express detectable levels of S-specific IgM, which may reflect differences in the time of sample collection between these studies.

Interestingly, we also detected differences related to immune subset composition among severe COVID-19 sub-cohorts, which were associated to the intensity of respiratory support defined as the type of oxygen, severity measured by PaO2:FiO2, and the presence of organ dysfunction defined by the WHO classification. In addition, a clinical debate persists as to why some severe COVID-19 patients get worse rapidly and require IMV while others display a more stable pathological course. In this context, we identified specific immune cell types that distinguished between severe COVID-19 patients under HFNC (displaying a more stable clinical course) versus patients who access rapidly to mechanical ventilation (Scores 6 and 7-9, respectively). These changes include increased frequencies of plasmablasts, monocytes, eosinophils, Th1 equivalents and underrepresentation of granzyme B^+^ producing CD8^+^ T cells in ventilated severe COVID-19 patients. How exactly these cell types contribute to promote or prevent the pathology remains to be fully elucidated.

Furthermore, we provide a longitudinal study of a small set of ICU patients which shows that innate and adaptive immune cells that were dysregulated during the onset of the pathology, including neutrophils, plasmablasts, activated CD4^+^ and CD8^+^ T cells, or that were underrepresented such as basophils, begin to regain normal proportions during recovery. Severe patients also synthesized markedly elevated levels of S-specific IgG during the entire time of the study, suggesting the generation of long-term immunity against the virus after infection. It remains to be clarified which of these immune alterations persists in time and have clinical significance in patients, emphasizing the need for more data emerging from longitudinal follow-ups of different cohorts of patients. Nevertheless, these data are consistent with very recent reports showing durable immune responses in recovered COVID-19 patients with heterogeneous degrees of disease severity after 8 months of infection (34,39).

Finally, our data is relevant considering that COVID-19 studies emerging from Latin America remain poorly represented in the literature. In addition, during this study, there have been drastic changes in the prevalence of SARS-CoV2 variants in Chile. The original variants present during the first half of 2020 have been replaced successively by VOCs alpha (B.1.1.7), and later by VOC gamma (P.1), and by VOI lambda (C.37) in the Chilean population (data from the national consortium of SARS-CoV-2 genome surveillance, https://auspice.cov2.cl/ncov/chile-global-2021-07-02). In Chile, the gamma and lambda variants have dominated the genomic landscape of SARS-CoV-2 in the first half of 2021, are also prevalent in many Latin American countries and remain conspicuously understudied. Further attention should be directed at immunological and clinical features associated with particular variants in local populations to detect potential effects of viral evolution on clinical outcomes.

## Data Availability

Not applicable

## Author contributions

Conceptualization: FO, MRB, ML

Data curation: PGG, VS, FO

Formal analysis: PGG, FO, LV, VS

Funding acquisition: FSO, RN, MH, FO, MRB, ML

Investigation: PGG, FT, LV, CF, CB, GL

Methodology: PGG, FT, GL, CF, CM, CB, RT, ET, LV, VS, MV, JPA, MN, AM, PM, MRa, VR, FO, MRB

Project administration: FT, ML

Resources: MA, MRB, FSO, RN, MH, FO, ML

Supervision: FO, ML, MRB Validation: PGG, LV

Visualization: PGG, FT, ML, MRB, FO

Writing-Original Draft Preparation: PGG, ML, MRB, FO

Writing-Review & Editing: PGG, FT, LV, VS, CF, CM, MA, VR, CB, MV, MRa, RT, JPA, MN, ET, MV, AM, PM, RN, GL, MH, FSO, ML, MRB, FO.

All authors accept responsibility to submit for publication.

## Acknowledgements

We thank patients and blood donors who consented to this study and the health team at the Hospital Clínico Universidad de Chile. We also thank members of the laboratory of cell therapy and experimental medicine for establishing appropriated conditions for experimental work in pandemic. We acknowledge the Millennium Institute on Immunology and Immunotherapy (IMII, ICM-ANID, ICN09_016) for access to critical flow cytometry equipment. We express our appreciation to the colleagues that belong to the Chilean SARs-CoV-2 Genome Surveillance Consortium and the Instituto de Salud Pública for their efforts to sequence viral genomes.

## Supplementary Figure Legends

**Supplementary Figure 1. COVID-19 study diagram. A**. In this study, 29 COVID-19 Moderate and 37 Severe COVID-19 patients, along with 28 convalescent patients and 28 Healthy controls were recruited from the Hospital Clínico Universidad de Chile for the study of the immunological landscape and disease evolution.

**Supplementary Figure 2. Gating strategies used for flow cytometric analyses of immune cell subsets. A**. General analysis of main immune populations found on peripheral blood mononuclear cells of patients (B cells, CD8 and CD4 T cells, Monocytes, DCs and basophils). **B**. Gating strategy used to examine CD8 and CD4^+^ T cell populations in peripheral blood of COVID-19 patients. **C**. Gating strategy used to identify different B cell populations (Transitional, non-transitional, plasmablasts, näive, IgD^-^CD27^-^and memory switch B cells) in fresh blood samples. **D**. Characterization of granulocytes and monocytes through flow cytometry analysis in fresh blood samples.

**Supplementary Figure 3. Granulocyte identification and Monocyte characterization from peripheral blood of COVID-19 patients. A**. Representative facs plots and frequency of monocyte populations of all subjects included in the study: healthy controls (HC), moderate (M), severe (S) and convalescent (C) COVID-19 patients. **B**. Representative histograms for HLA-DR and CD86 expression assessed in the different monocyte subsets (conventional, intermediate and non-conventional monocytes). **C**. Representative histograms for PD-L1 expression determined in neutrophils and monocytes from all conditions studied (healthy controls (HC), moderate (M), severe (S) and convalescent (C) COVID-19 patients).

**Supplementary Figure 4. Identification of cellular immune components in peripheral blood of COVID-19 patients. A**. Representative facs plots and frequency of transitional and non-transitional B cell populations for all conditions included in the study: healthy controls (HC), moderate (M), severe (S) and convalescent (C) COVID-19 patients. **B**. Representative facs plots and frequency of näive, IgD^-^CD27^-^and memory switched B cells identified in all 4 subjects studied. **C**. Representative facs plots and frequency of CCR6^+^CXCR3^-^Th17^-^equivalents characterized in all 4 conditions studied (healthy controls (HC), moderate (M), severe (S) and convalescent (C) COVID-19 patients).

