## Supplementary Figures for "Dysregulated immune responses in COVID-19 patients correlating with disease severity and invasive oxygen requirements"

**Supplementary Figure 1**

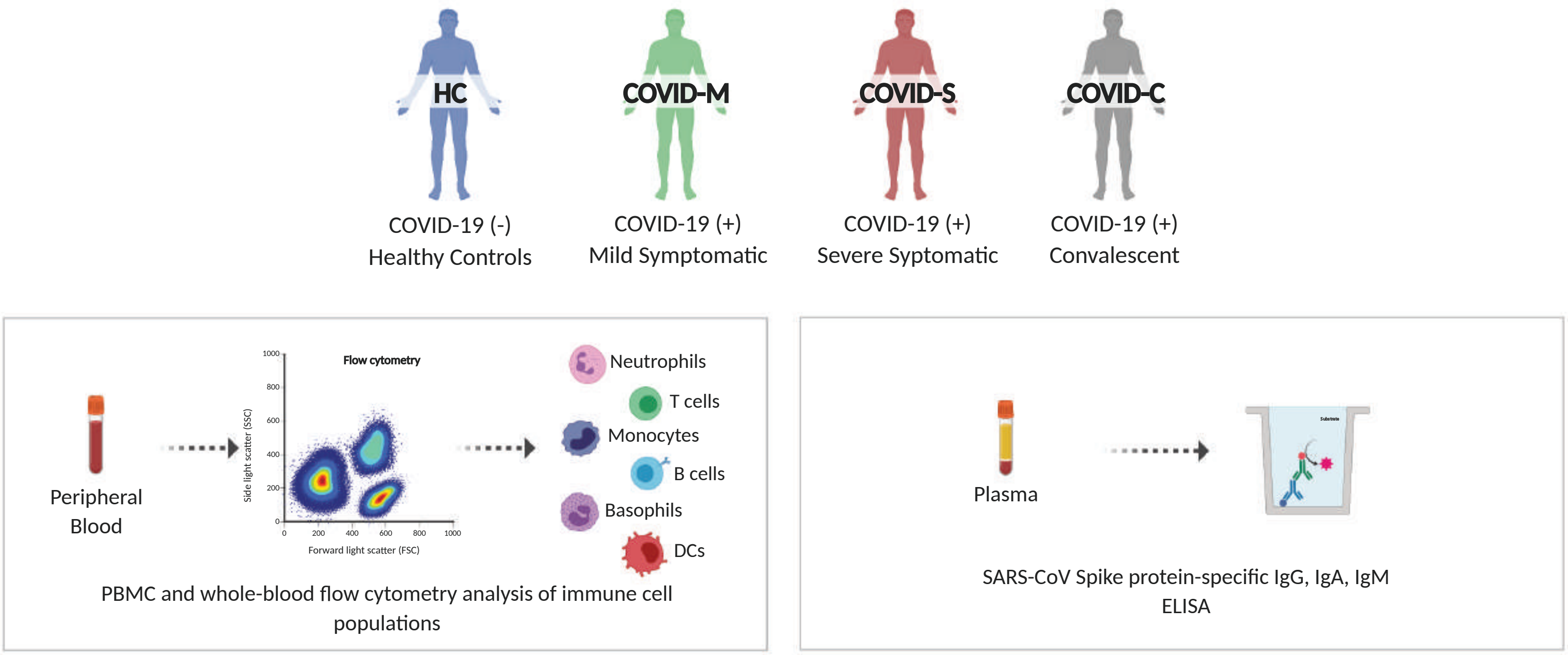

Supplementary Figure 2

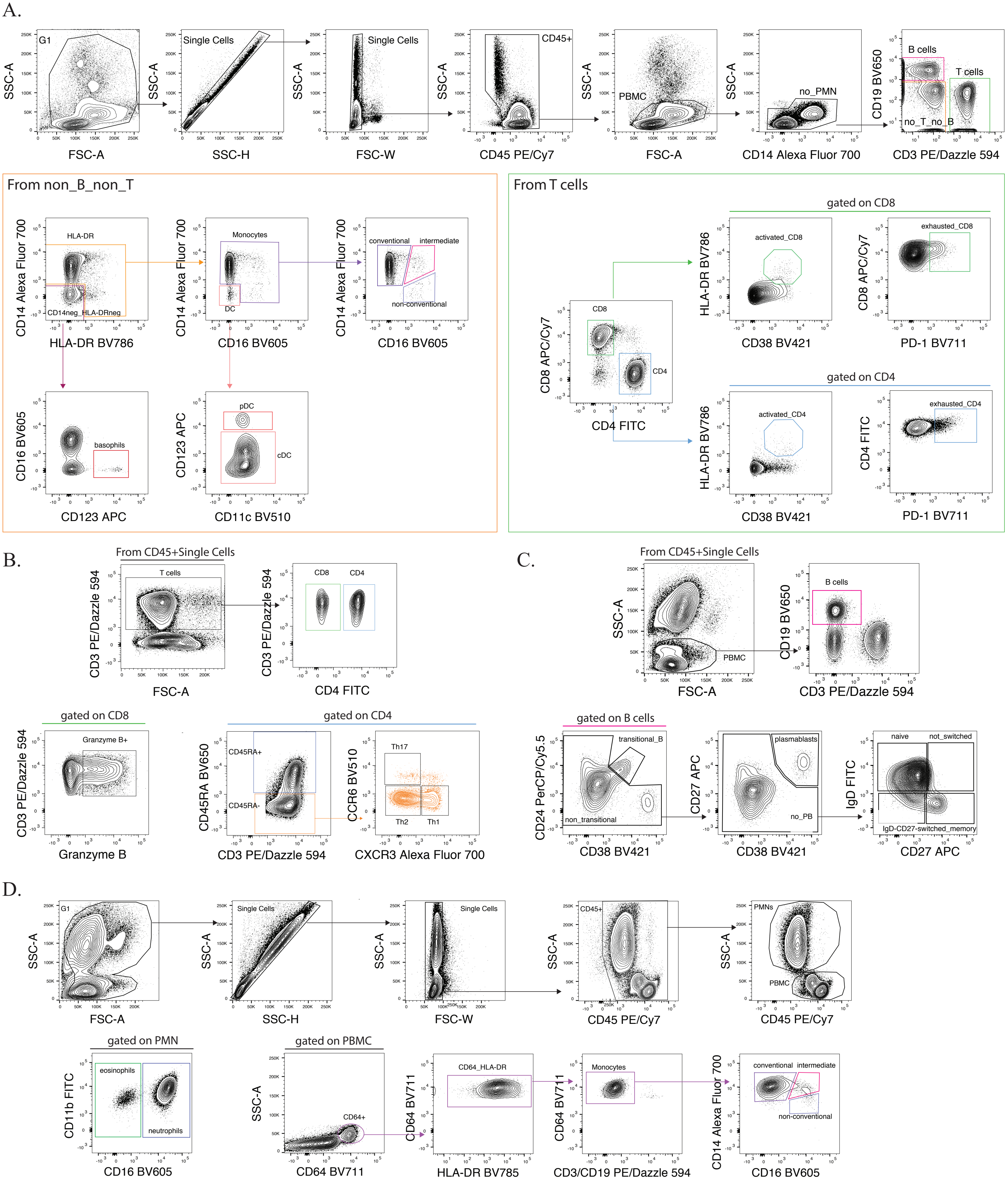

### Supplementary Figure 3

**A.**

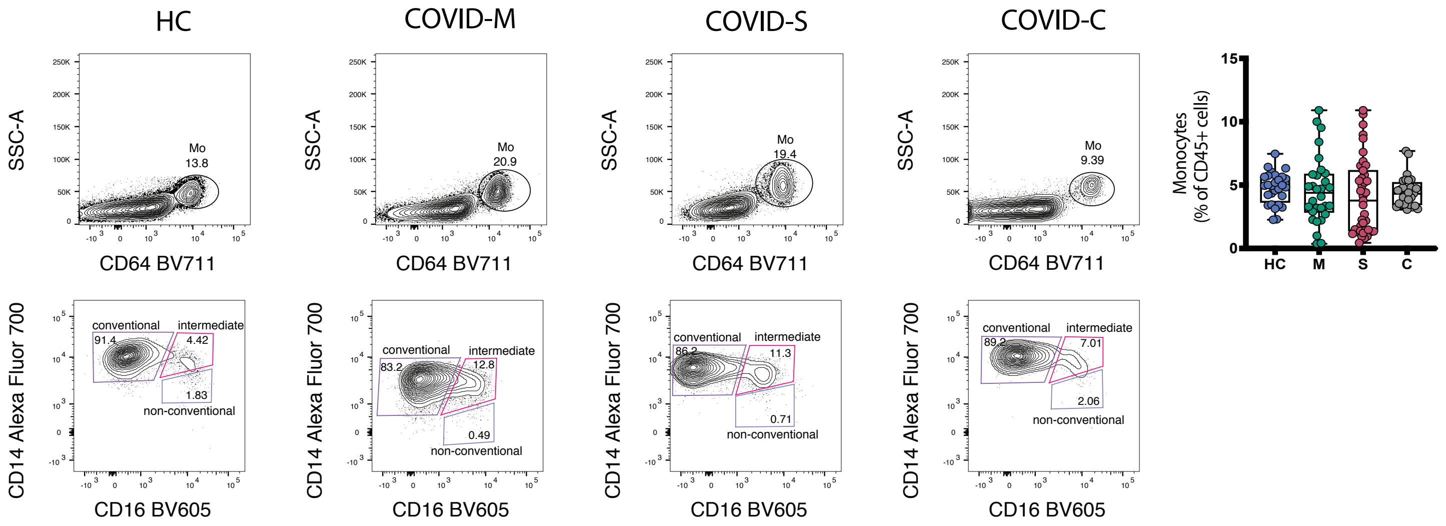


**B.**

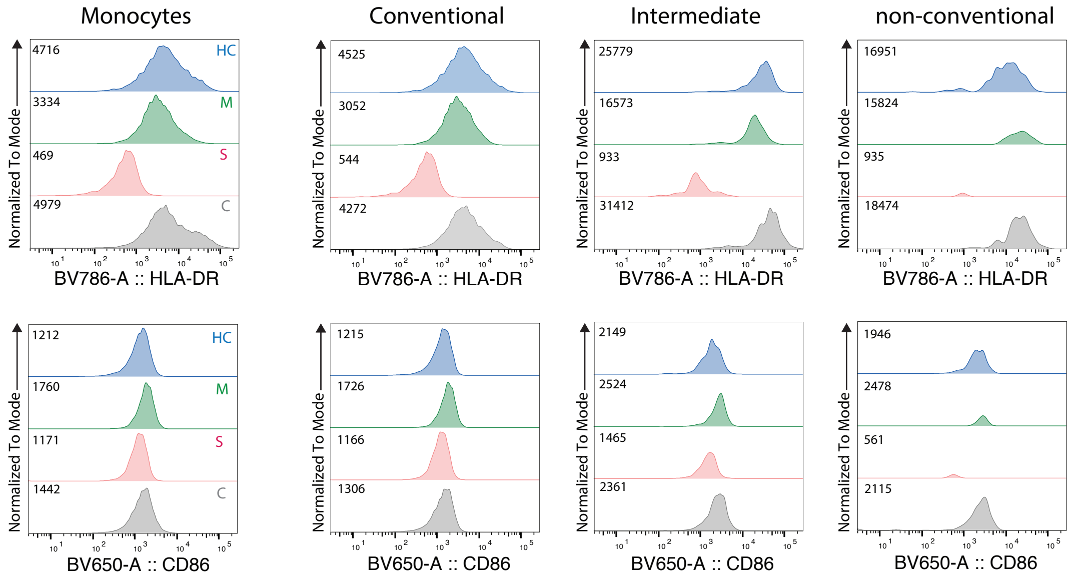


**C.**

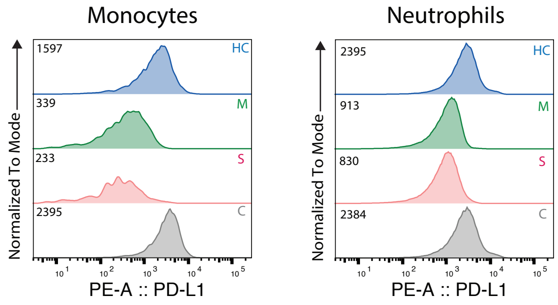

#### Supplementary Figure 4

**A.**

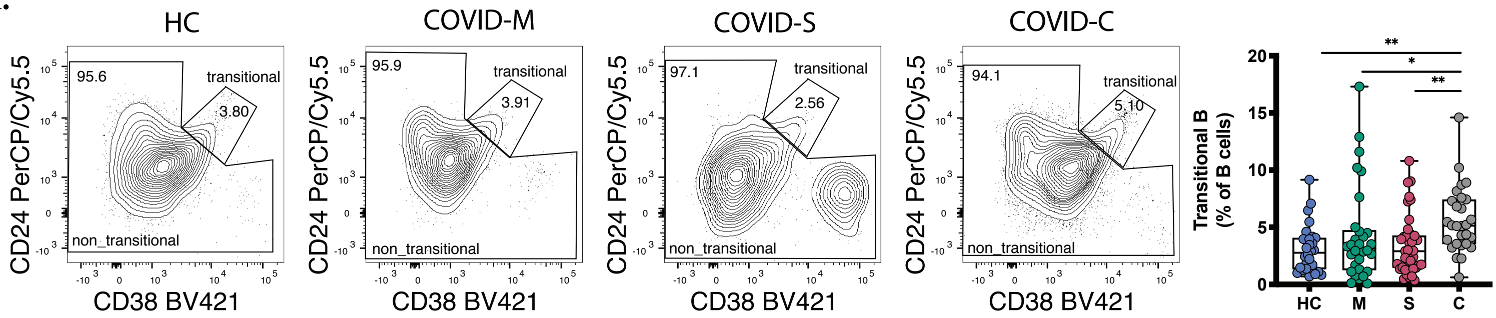


**B.**

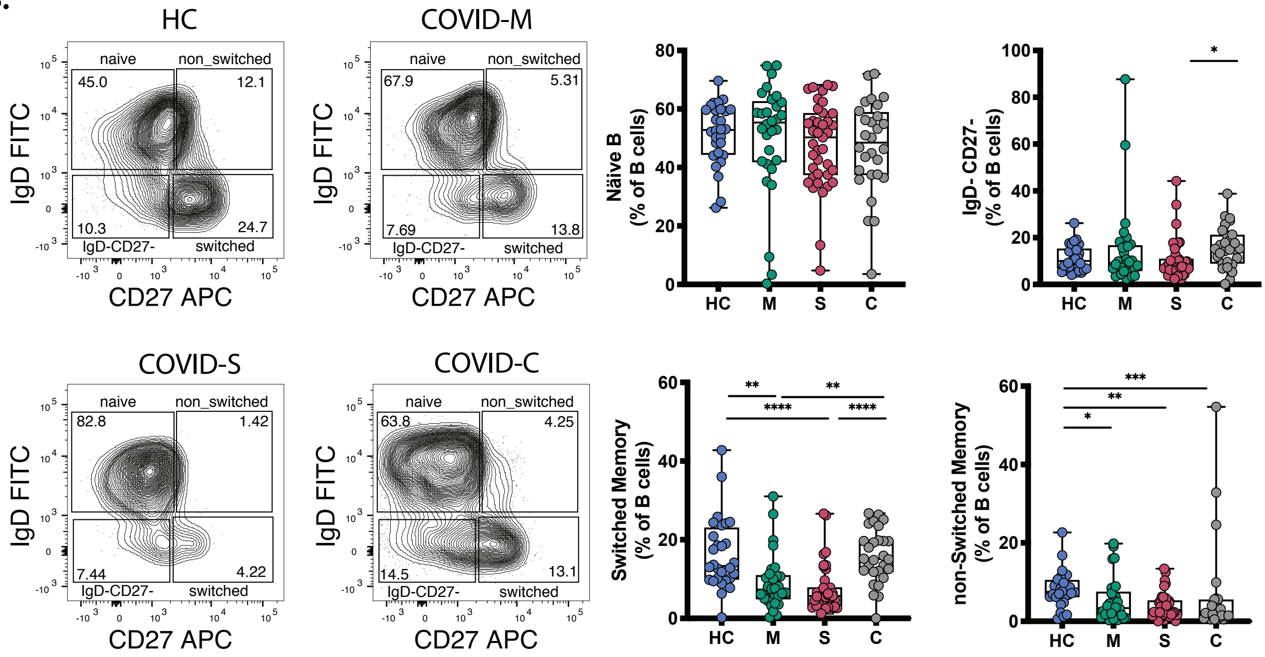
